# Incremental cost of pre- and post-exposure prophylaxis service provision via an online pharmacy in Kenya

**DOI:** 10.1101/2025.03.14.25324000

**Authors:** Yilin Chen, Michalina A Montaño, Paulami Naik, Nicholas Thuo, Catherine Kiptinness, Maeve Rafferty, Andy Stergachis, Melissa Latigo Mugambi, Kenneth Ngure, Katrina F. Ortblad, Monisha Sharma

## Abstract

**Background:** Online pharmacy HIV pre- and post-exposure prophylaxis (PrEP/PEP) provision is a novel strategy to expand HIV prevention coverage. In the ePrEP pilot study, we found online pharmacy PrEP/PEP was feasible and reached populations at HIV risk in Kenya. However, program costs data are lacking.

**Methods:** We conducted a costing within the ePrEP pilot study in Nairobi from 11/01/2022-12/29/2023. We obtained costs from expense reports and conducted time-and-motion observations and staff interviews. We estimated total and unit costs in the first year of implementation, cost per client and per PrEP client-month (2023 US Dollars (USD)).

**Results:** Overall, 229 clients initiated PrEP (507 months of PrEP coverage) and 1320 initiated PEP. Based on observed program volume, annual financial cost was $109,945 USD (PrEP: $19,456; PEP: $90,489). Cost per client was higher for PrEP than PEP ($85 vs $68.6), and cost per PrEP client-month was $38 (mean duration: 2.2 months). Main drivers of financial costs were courier-delivery of HIV testing kits and drugs (PrEP: 50.6%; PEP: 40.5%), demand generation (PrEP: 25.9%; PEP: 32.1%), and equipment, system development, and utilities (PrEP: 9.3%; PEP: 9.8%). Assuming a scaled-up client volume of 2500 (PrEP: 370; PEP: 2130) reduced per-client financial costs for PrEP ($65.5) and PEP ($56) and cost per PrEP client-month ($29.6).

**Conclusions:** Costs of online PrEP/PEP provision is likely higher than clinic-based PrEP. Implementing cost sharing models including charging clients for HIV testing and optimizing courier delivery routes can increase program efficiencies. Our cost estimates can inform economic evaluations of online PrEP/PEP delivery.

## INTRODUCTION

Despite substantial strides in HIV treatment and prevention, HIV remains a leading cause of morbidity and mortality, disproportionately impacting Eastern and Southern Africa (ESA) [1]. Oral pre-exposure prophylaxis (hereafter referred to as PrEP) is highly effective at preventing HIV acquisition and has been recommended by the World Health Organization (WHO) since 2015 for use among persons with HIV risk indication [2,3]. In 2017, the Kenyan Ministry of Health implemented PrEP service delivery within government clinics as part of the national HIV prevention strategy. However, uptake and persistence of clinic-based PrEP remains low; barriers include long travel and wait times, stigma, privacy issues, and limited clinic hours [4,5]. PrEP provision in community-based settings including pharmacies has been shown to achieve higher PrEP uptake than facility settings [6,7]. Similarly, post-exposure prophylaxis (PEP) is a promising biomedical prevention that is effective at reducing HIV acquisition when used within 72 hours after a potential exposure [8]. Availability of PEP is generally limited to clinic settings but increasing availability via community-based channels is projected to be an efficient strategy to reduce HIV incidence [9].

Leveraging online pharmacies for PrEP/PEP provision is a novel strategy that could expand HIV prevention coverage by providing convenient and discreet services in the community. Online pharmacies offer direct to consumer delivery of medications, health products, health information, and remote clinical consultations [10]. Telehealth platforms are rapidly expanding in ESA with growing internet availability and cell phone use, and expansion was expedited by the COVID-19 pandemic [11–13]. In a recently completed pilot study (ePrEP Kenya), our team tested the first model of online pharmacy-based PrEP/PEP service delivery in Africa in collaboration with MYDAWA, Kenya’s first licensed online pharmacy [14]. The model entailed clients ordering a courier-delivered HIV self-test (HIVST) or provider administered HIV test via online pharmacy (to verify HIV-negative status), completing a remote consultation with a clinical provider to ensure PrEP/PEP is not contraindicated, receiving drug delivery to one’s home or a convenient location, and having access to virtual user support. We found that online pharmacy-based PrEP/PEP was feasible, acceptable, in demand, and reached populations at HIV risk who are distinct from those accessing clinic-based PrEP [15,16].

Estimating the incremental costs of online pharmacy PrEP/PEP provision is essential to informing strategies for sustainable scale up. Cost estimates can also serve as inputs in economic evaluations to assess cost-effectiveness and budget impact of prevention programs. Online service delivery of biomedical prevention will require partnerships between private sector and government agencies and evaluations of potential cost sharing models to provide drugs and HIV testing. We sought to estimate the incremental costs and cost drivers of providing PrEP and PEP via online pharmacy in Nairobi, Kenya. While previous studies have estimated the costs of clinic-based PrEP service delivery [17–21], to our knowledge, this is the first study to estimate the costs of online PrEP services and among the first to evaluate the cost of PEP provision.

## Methods

### Study Setting

We conducted cost data collection within the ePrEP Kenya pilot in Nairobi. The study protocol has been previously published [22]. Briefly, the pilot was conducted from November 2022 to December 2023 in collaboration with MYDAWA, Kenya’s first licensed online pharmacy (https://mydawa.com) [10]. MYDAWA sells health products (including those related to sexual health and contraception), prescriptions, and over-the-counter medicines which are directly delivered to consumers.

### Intervention

Online PrEP services were advertised on the MYDAWA website and via outreach events. Chatbot and call centers were set up to answer potential clients’ questions. Study inclusion criteria were: ≥18 years old, self-identified at risk of HIV, no medical contraindications, smart phone or computer access, and having a delivery address within Nairobi County. Interested individuals booked a telehealth visit with a trained clinical officer via the MYDAWA online platform. Visits were generally completed on participants’ phones (using video or voice call); providers assessed PrEP eligibility and provided counseling on HIV testing, PrEP/PEP, adherence, and risk reduction. During the telehealth visits, providers used a prescribing checklist to assess client’s eligibility for online PrEP and PEP including sexual behavior and HIV exposure in the past 72 hours. Clients were screened for acute HIV infection and medical conditions that might contraindicate PrEP/PEP (**Figure S1**).

At the end of the telehealth visit, clinicians transferred conditional prescription information to the warehouse using the MYDAWA app (pending confirmation of HIV-negative status). Warehouse staff prepared pick-up instructions and dispatched orders though a “Get boda” app that assigns orders to available riders and enables clients to track delivery time. Eligible clients were directed to purchase their choice of either an HIV self-test (HIVST) or counselor-administered rapid diagnostic test (RDT) for a respective $3 or $2 USD fee. HIVSTs were delivered by pharmtech riders for a $1 USD delivery fee to clients’ preferred location.

Clients then used the HIVST and uploaded an image of their test result via a MYDAWA secure platform; a clinician confirmed HIV-negative status at the time of delivery. The pilot study incorporated the use of artificial intelligence (AI) to assist the clinical officers in reading HIVST results. AI provided an interpretation of HIVST results which was then verified by the clinical officer. The pharmtech rider delivered a 30-day prescription of PrEP or PEP to clients after eligibility were determined. Pre-testing consultation on use of HIVST and post-testing consultation on medication usage and safety were provided by the pharmtech during the delivery process. A brief follow-up call was conducted one week post PrEP/PEP initiation. Clients interested in continuing PrEP at one-month post-initiation completed HIV testing and received a 90-day PrEP supply once continued eligibility was confirmed; clients could continue to receive PrEP refills every 3 months thereafter. PEP clients were scheduled to complete an additional visit at 28 days post PEP initiation in which clinical officers provided counseling about transitioning from PEP to PrEP. For the majority of clients, pharmtech riders delivered HIVST kits and PrEP/PEP medications in a single trip; they waited for HIV test results and confirmation of negative status/eligibility before dispensing drugs.

The research protocol was reviewed and approved by the Scientific and Ethics Review Unit at the Kenya Medical Research Institute (Nairobi, Kenya) and was granted an exempt status by the Institutional Review Board at the University of Washington (Seattle, USA). Participants completed informed consent prior to receiving services.

### Cost data collection

We conducted micro-costing following Global Health Cost Consortium Reference Case guidelines to estimate the incremental economic and financial cost of online PrEP/PEP delivery from the provider perspective [23]. The financial costs captured only costs required to provide online PrEP/PEP services, and therefore most closely represented real-world costs of PrEP/PEP provision by an online delivery service provider. Economic costs included the opportunity costs of donating goods and existing overhead including drug and vehicle costs for delivering HIVST and PrEP/PEP drugs. Start-up costs included initial microplanning and training activities, initial demand generation, and system development (HIVST uploading and interpretation system, telehealth, chatbot, and call center solution). Recurrent costs included personnel salaries and benefits, ongoing microplanning and refresh training, continued demand generation and marketing, clinical consultation, delivery of HIVST and drugs, user support, equipment, fuel, and utilities (**Table S1**).

We estimated personnel time costs (e.g., clinical officer, pharmtech rider, warehouse staff, and call center staff) using time-and-motion studies to estimate average time providers spent in on different activities (in minutes), multiplied by personnel cost per minute (accounting for salary and benefits and hours worked/year). Time and motion observations were conducted midway through the pilot to capture personnel time when the program was efficiently running. For other supporting personnel (e.g., project coordinator, administrator), we proportionally allocated their salary and benefits based on the percentage of time dedicated to online PrEP/PEP delivery. For supplies and commodities, we observed resource use by activity during time-and-motion and multiplied quantity used by unit costs obtained from expense reports, program budgets, or centralized price lists. Drug costs include 200 mg of emtricitabine and 300 mg of tenofovir disoproxil fumarate (Truvada: $4.28 USD per 30 days) for PrEP and 50mg of Dolutegravir, 300mg of Lamivudine, and 300mg of Tenofovir disoproxil (Acriptega: $6.03 USD per month) for PEP, respectively [24]. The Kenya Ministry of Health provided PrEP and PEP drugs for free for the pilot and clients were not charged for drugs, therefore drugs were included as economic costs only. We excluded research activities as they would not be performed in PrEP/PEP services. Key PrEP/PEP delivery components costs are listed in **Table S2;Table S3** shows model assumptions.

Costs were collected in Kenyan Shillings (KES) and converted to 2023 United States Dollars ($1 USD = 156 KES). We annualized start-up and capital costs over the average useful life (five years) using a 3% annual discount rate [25]. The analysis was conducted in Excel 2018 (Microsoft, Redmond, USA). Details on costing methods and the Excel file used for analysis are provided in the Supplementary Information.

### Program volume and cost analysis

We utilized study monitoring data to estimate number of individuals who initiated PrEP/PEP and continued PrEP over a one-year pilot period. We estimated number of annual PrEP/PEP telehealth visits by assuming that all PrEP clients had one telehealth visit at initiation, followed by a 1-week follow-up call. The number of continuation visits was determined based on observed PrEP continuation at 1, 4, 7, and 10 months. PEP clients were scheduled to have one telehealth visit at PEP initiation, a follow-up call at 1-week, and another visit at 28 days. We assumed 10% of PEP clients completed follow-up call and visits based on staff interviews. The annual cost for online PrEP/PEP was calculated by multiplying the number of initiation and continuation visits by their respective average unit costs and summing the total. We calculated cost per PrEP client, PEP client, client-month of PrEP dispensed, PrEP initiation, and PrEP continuation as follows:

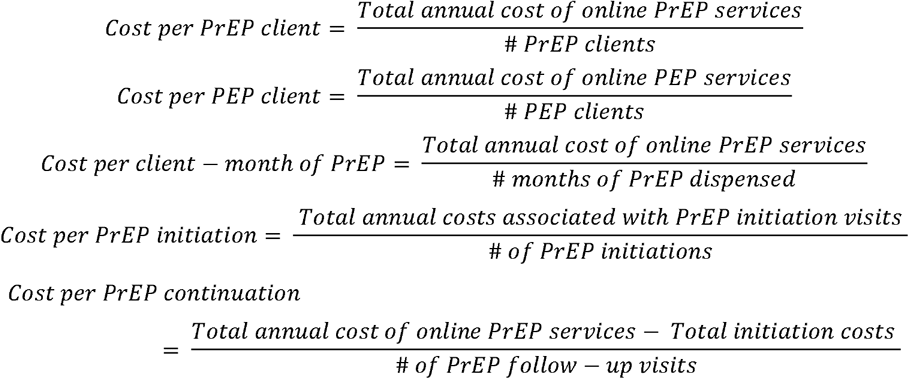

### Scenario analyses

We evaluated the cost of different delivery scenarios. In one scenario, we assumed introduction of a novel $1 USD HIVST kit into the market. This was motivated by the partnership between the Clinton Health Access Initiative, MedAccess, and Wondfo Biotech Company which will likely introduce a HIV self-test for $1 USD in the public sector in low- and middle-income countries [26]. Separately, we evaluated program costs under a range of plausible client volumes up to 2500, the estimated program capacity at scale without requiring additional resources. We fixed the proportion of PrEP and PEP clients in this analysis (PrEP: 15%; PEP: 85%) to match the pilot study. In the base case, we assumed that only clinical officers interpreted the HIVST results to reflect real-world practice, making findings more applicable to other settings. In a scenario analysis, we assessed the impact of including the AI-enabled HIVST platform. This scenario included costs of AI development, system integration and set up, and AI interpretation. Additionally, we varied the duration of PrEP initiation and follow-up visits by ±30% to assess the impact of PrEP visit times on costs. Lastly, we explored a scenario of task-shifting PrEP/PEP provision from highly paid pharmtech riders to lower cadre workers who had a 50% lower salary.

## RESULTS

### Program volume

During the one-year pilot, 229 individuals initiated PrEP, accruing 507 total months of PrEP coverage (mean duration: 2.2 months, median: 1, IQR:1, 2.1); additionally, there were 1,320 online PEP initiations.

### Financial and economic costs of online PrEP/PEP provision

Utilizing the annual PEP and PrEP client volume observed in the pilot, we estimated the annual financial cost of online PrEP/PEP delivery as $109,945 USD, with $90,489 USD of costs allocated to PEP services and $19,456 USD allocated to PrEP. The estimated annual economic cost of online PrEP/PEP provision was $31,212 USD for PrEP and $98,467 USD for PEP services (**Table 1**). Annual cost by category is displayed in **Figure S2**. The cost per PrEP client was $85 USD (financial) and $136.3 USD (economic) and cost per client-month of PrEP was $38 USD (financial) and $61.6 USD (economic). The per-client cost for PrEP initiation (financial: $43 USD; economic: $75.2 USD) was greater than that of PrEP continuation (financial: $26.6 USD; economic: $59 USD). For PEP, the cost per client was $68.6 USD (financial) and $74.6 USD (economic) (**Table 1**).

**Table 1.** Total and unit costs of online PrEP and PEP provision in Kenya (2023 USD)

| Cost Category | Total Cost: PrEP<br>(n=229) |  | Total Cost: PEP<br>(n=1320) |  | Unit Cost: PrEP<br>(n=229) |  | Unit Cost: PEP<br>(n=1320) |  |
| --- | --- | --- | --- | --- | --- | --- | --- | --- |
|  | Economic | Financial | Economic | Financial | Economic | Financial | Economic | Financial |
| Salaries and Benefits | \$1,138 | \$1,138 | \$6,559 | \$6,559 | \$5.0 | \$5.0 | \$5.0 | \$5.0 |
| Microplanning and Training | \$277 | \$277 | \$1,596 | \$1,596 | \$1.2 | \$1.2 | \$1.2 | \$1.2 |
| Demand Generation and Marketing | \$5,034 | \$5,034 | \$29,017 | \$29,017 | \$22.0 | \$22.0 | \$22.0 | \$22.0 |
| Clinical Consultation | \$1,800* | \$1,800* | \$8,832<sup>†</sup> | \$8,832<sup>†</sup> | \$7.9 | \$7.9 | \$6.7 | \$6.7 |
| Delivery of HIVST and Drugs | \$9,844 | \$9,844 | \$36,628 | \$36,628 | \$43.0 | \$43.0 | \$27.7 | \$27.7 |
| User Support | \$104 | \$104 | \$599 | \$599 | \$0.5 | \$0.5 | \$0.5 | \$0.5 |
| Equipment, System Development, Utilities, and Fuel | \$1,259 | \$1,259 | \$7,258 | \$7,258 | \$5.5 | \$5.5 | \$5.5 | \$5.5 |
| Drug Costs | \$11,753 | - | \$7,962 | - | \$51.3 | \$0.0 | \$6.0 | \$0.0 |
| Vehicles | \$3 | - | \$16 | - | \$0.01 | \$0.0 | \$0.01 | \$0.0 |
| **Total Cost** | **\$31,212** | **\$19,456** | **\$98,467** | **\$90,489** | | | | |
| **Cost Per Client** | **\$136.3** | **\$85** | **\$74.6** | **\$68.6** | | | | |
| **Cost Per PrEP Client Month** | **\$61.6** | **\$38.4** | | | | | | |
| **Cost Per PrEP Initiation** | **\$75.2** | **\$43** | | | | | | |
| **Cost Per PrEP Continuation** | **\$59** | **\$26.6** | | | | | | |

\*For PrEP, this includes an initiation visit, a brief follow-up call at 1 week, and PrEP continuation visits scheduled at 1-, 4-, 7-, and 10-months following initiation, respectively.
<sup>†</sup>For PEP, this includes an initiation visit, a follow-up call at 1 week, and an additional visit at the 28th day after initiation.

The main drivers of financial cost were courier delivery of commodities (HIVST kits and drugs; 50.6% for PrEP, 40.5% for PEP), followed by demand generation and marketing (25.9% for PrEP, 32.1% for PEP), and equipment, system development, and utilities (6.5% for PrEP, 8.0% for PEP). The main drivers of PrEP economic cost were drugs (37.7%), followed by delivery (31.5%), and demand generation and marketing (16.1%). In contrast, drug was not a main driver of PEP economic costs (8.1%), but rather the delivery of HIVST and drugs (37.2%), and demand generation and marketing (29.5%). Clinical consultation costs were similar across PrEP and PEP ($7.9 vs $6.7 per client, respectively) as while costs of delivering HIVST and drugs were higher for PrEP clients due to the greater number of deliveries needed for PrEP initiation and continuation ($43 per PrEP client and $27.7 per PEP client) (**Figure 1**). The annual costs for delivery of HIVST and drugs for both PrEP/PEP orders was $46,472 USD (PrEP: $9,844 USD; PEP: $36,628 USD) (**Figure S2**), which constituted the largest proportion of all personnel time spending in the pilot project (80%).

**Figure 1.**
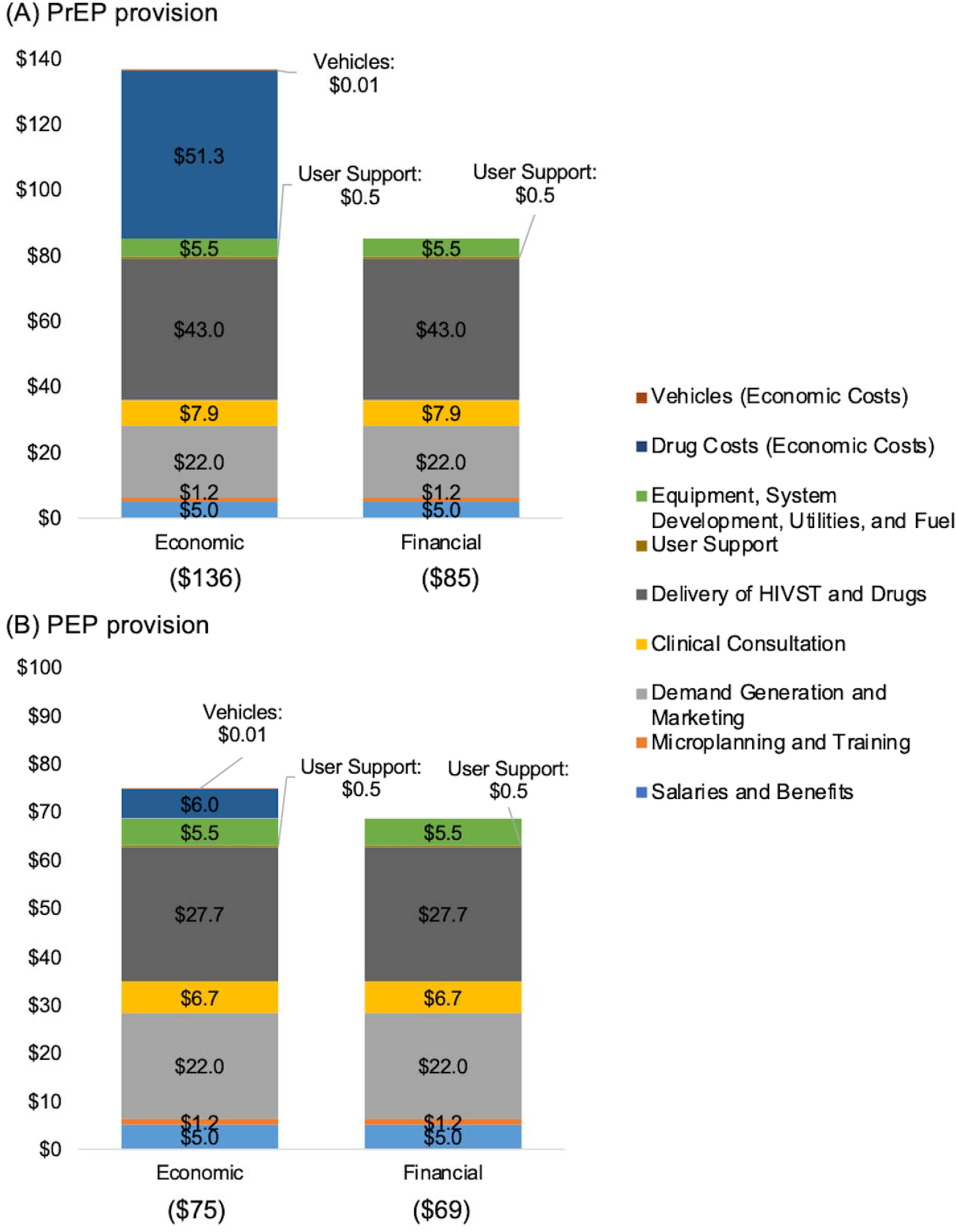
Costs of online PrEP/PEP provision per client (2023 USD)

### Time-and-motion observations

We conducted 12 time-and-motion observations, including 7 PEP and 1 PrEP visits with clinical officers and 4 observations for order preparation and dispatch at the warehouse. Clinical officers spent a mean of 18 minutes on PEP initiation visits (standard deviation [SD] 10 minutes), which entailed eligibility assessment, counseling, and medication prescribing. PEP follow-up visits were shorter than PEP initiation visits, lasting an average of 6 minutes (SD: 2.1 minutes), and consisted of clinical officers providing counseling and assessing any potential PrEP safety issues or adverse events. We observed one PrEP initiation visit which lasted 11 minutes and included: assessment of HIV risk and health history, PrEP consultation, prescription, and answering client questions (**Table 2**). We assumed the duration of PrEP follow-up visits were equivalent to that of PEP follow-up visits based on staff interviews. Time and motion observations and assumptions were confirmed in staff interviews to more accurately reflect average personnel time needed for visits.

**Table 2.** Time (minutes) for clinical service delivery components estimated from time-and-motion studies and unit costs (USD 2023)

| Activity | Minutes | Hourly Salary | Total Cost |
| --- | --- | --- | --- |
| ***Clinical consultation (per visit)*** |  |  |  |
| PEP initiation | 18 | 17 | \$4.98 |
| PEP 28-day follow-up | 6 | 17 | \$1.66 |
| PEP 1-week follow-up call | 6 | 17 | \$1.66 |
| PrEP initiation | 11 | 17 | \$3.04 |
| PrEP 1-week follow-up call | 6 | 17 | \$1.66 |
| PrEP continuation follow-up | 6 | 17 | \$1.66 |
| **Subtotal** | | | **\$14.65** |
| ***Delivery of HIVST and drugs*** |  |  |  |
| Warehouse preparation per delivery | 22 | 8 | \$3.04 |
| Pharmtech rider per delivery | 90 | 8 | \$12.10 |
| **Subtotal** | | | **\$15.14** |
| ***User support*** |  |  |  |
| Call center support per call | 1.3 | 6 | \$0.12 |
| **Subtotal** | | | **\$0.12** |

On average, preparing and dispatching prescriptions at the warehouse required 22 minutes (SD: 1.3 minutes); warehouse staff spent 63% of their time pre-processing orders. Pharmtech rider time needed to complete a delivery varied considerably depending on delivery distance. According to pharmtech riders, deliveries during one trip required 90 minutes, which included pre-testing counseling, HIV testing and uploading of the test result, post-testing counseling, and driving time (**Table 2**). This translates to $15.1 USD per delivery of HIVST and drugs.

### Scenario analysis

Assuming availability of a $1 USD HIVST resulted in lower annual financial cost, with greater reduction for PrEP (−2% change) than PEP services (−1.6% change), translating to savings of $1.7 USD PrEP client and $1.1 USD for PEP. The financial cost per PrEP client-month was reduced from $38.4 USD to $37.6 USD (−2.1%) compared to the base case with this scenario (**Table S4**).

Assuming expanded volume of online PrEP/PEP clients to 2500 (PrEP: 370, PEP: 2130) was projected to reduce cost per client to: $65.5 USD (financial; -23% from the base case) and $116.9 USD (economic; -14.2%) for PrEP services and $56 USD (financial; -18.3%) and $62.1 USD (economic; -16.8%) for PEP services. The estimated cost per PrEP client month was $28.3 USD (financial) and $51.5 USD (economic) (**Figure S3**). In a scenario assuming more PrEP clients (500 instead of 229 in the base case) and 2000 PEP clients, the financial and economic cost per PrEP client further decreased to $62.7 (−26.2%) and $114.1 (−16.3%), resulting in the financial and economic cost per client-month of $28.3 and $51.5, respectively.

Including AI system development and integration costs in the model resulted in increased financial and economic costs per client for PrEP and PEP. The estimated cost per PrEP client month was $41.4 USD (financial) and $64.4 USD (economic) (**Table S5**).

Additionally, increasing or reducing the PrEP initiation and follow-up visit time by 30% was estimated to have a minimal impact on the program costs (**Table S6**). Finally, Assuming PrEP/PEP delivery was conducted by lower-cadre workers earning 50% less than current pharmtech riders, the per-client cost decreased by $10.3 for PrEP and $6.7 for PEP in both economic and financial cost estimates (**Table S7**).

## DISCUSSION

To our knowledge, this is the first analysis to estimate the financial and economic costs of providing PEP and PrEP via an online pharmacy in Eastern and Southern Africa. We estimated financial and economic costs of $85 and $136.3 per PrEP client, and $68.6 and $74.6 per PEP client. The financial and economic cost per PrEP client-month was $43 and $75, respectively. Assuming a scaled-up program with 2500 clients lowered per client costs to $65.5 USD (financial) and $116.9 USD (economic) for PrEP and $56 USD (financial) and $62.1 USD (economic) for PEP. The largest component of PEP economic costs was personnel time costs for delivery of the HIVST and PEP drugs, followed by demand generation and marketing costs. Conversely, PrEP medication constituted the largest component of PrEP economic costs, followed by HIVST and drug delivery, and demand generation. Notably, personnel time and associated clinical consultation costs were low compared to other categories. The PrEP financial cost per client was higher than that of PEP, largely due to additional drugs and delivery associated with PrEP continuation visits. The cost per PrEP initiation was higher that of PrEP continuation due to longer clinical consultation in the initiation visit. Delivery costs were particularly high, accounting for approximately half of the financial costs for PrEP and 40% for PEP, due to the high salary cost of the pharmtech rider. Additionally, a single client order was fulfilled per delivery. Task shifting to lower cadre riders and batching deliveries can greatly reduce these costs.

Several studies have examined the cost of PrEP delivery via maternal and child health (MCH), HIV care, and family planning clinics in Kenya. One study estimated the economic cost of PrEP provision in public HIV care clinics as $21.3 per client-month (2019 USD), with 33% and 43% of the cost attributable to PrEP medication and personnel, respectively [19]. Another study estimated the economic cost of PrEP provision through MCH and family planning clinics as $26.52 (2017 USD) per client-month, with personnel and PrEP drugs contributing 43% and 25% of program costs, respectively [20]. We found economic cost of delivering PrEP via an online pharmacy were higher than clinic-based models in the literature, likely due to high costs of demand generation and delivering HIVST and drugs ($22 and $43 per PrEP client). The per client cost of clinical consultation was low for both PrEP and PEP services ($7.9 and $6.7).

Expanding the program to more clients will substantially reduce the cost per client and make the costs comparable to clinic-delivered models, as shown in our scenario analyses. Consistent with the literature, we found that drug and personnel costs were the main cost drivers of PrEP; however, in our model, most of the delivery costs are attributable to personnel time of pharmtech drivers, rather than clinical consultations.

Online delivery of PrEP and PEP is a new service in Kenya, and it is likely that this program will undergo future modifications that may impact the cost of services. Our scenario analyses show that higher client volumes would result in substantial decreased service cost, similar to other studies [17,18,27]. Future studies assessing online PrEP delivery costs are needed as the program moves beyond the initial phase. Cost of demand generation will likely decrease over time. Start-up expenses, such as system development, will have less of an impact after the program scales up. Moreover, further streamlining of the dispatch and delivery process can enhance efficiency, such as optimizing delivery routes to deliver products to multiple clients or in bundled packages. Relatedly, it will be important to monitor PrEP discontinuation. Previously clinic-based PrEP costing studies found that longer PrEP continuation decreases the per client-month cost of PrEP delivery due to the higher relative costs of PrEP initiation visits compared to continuation visits [19,27].

The majority of the total annual costs of PrEP/PEP implementation were driven by costs of PEP provision due to the higher assumed volume of PEP clients. Empirical data from the ePrEP/PEP pilot study found that PEP uptake was 7-times higher than PrEP uptake despite lack of demand generation for PEP services [15]. This may be due to individuals with a recent potential HIV exposure having higher motivation to seek prevention services compared to those who are seeking PrEP for potential exposures and are less sure that an exposure will occur.

These findings may highlight a need for short-term event driven services which are easier to fit into individual lives compared to long-term PrEP use. This pilot study was among the first to assess community demand for PEP so future studies are needed to assess comparative demand for PEP vs PrEP.

Our results have several limitations. First, we only observed one PrEP initiation visit, and no PrEP follow-up visits due to infrequent PrEP initiations and short data collection period, which may impact estimate accuracy. However, our observed estimates were confirmed by staff interviews and we used staff-reported time estimates for PrEP follow-up visits. In addition, we assessed online PrEP/PEP provision only in Nairobi County, which may not be generalizable to other geographic regions and other online pharmacies. Additionally, delivery time costs were substantial which likely vary by setting, particularly for rural regions where delivery time may be greater. Finally, we assumed that recurrent demand generation activities occur once every year to keep up with current trends which may occur less frequently over time.

Our cost estimates offer valuable insights for policymakers and private sector stakeholders considering the implementation of online pharmacy-based PrEP and PEP service provision in Kenya and similar settings. Personnel time costs for delivering HIVST and drugs represent approximately half of the incremental financial costs associated with online PrEP and PEP provisions. Efforts to improve the efficiency of online PrEP/PEP services should prioritize optimizing service delivery strategies and reducing expenditures on demand generation activities. Findings can inform cost-effectiveness and budget impact analyses to evaluate the affordability of leveraging online pharmacies for HIV prevention in Kenya.

## Data Availability

All data produced in the present study are available upon reasonable request to the authors

## Acknowledgements

We are grateful to the participants and staff of the online pharmacy PrEP/PEP pilot study. We thank MYDAWA staff including Maeve Rafferty, and Lisbet Charana for their support and assistance with data collection.

