## Supplemental Materials for "Incremental cost of pre- and post-exposure prophylaxis service provision via an online pharmacy in Kenya"

**Supplementary Materials**

Table S1. Input costs of key PrEP/PEP delivery components (2023 USD)

Table S2. List of key model assumptions

Table S3. Cost categories, description, and data sources for online PrEP/PEP provision

Table S4. Scenario analysis of using $1 HIVST kit

Table S5. Scenario analysis of including AI HIV testing system

Table S6. Scenario analysis of varying duration of PrEP initiation and continuation visits

Table S7. Scenario analysis of using lower-cadre pharmtech workers

Figure S1. The proposed care pathway for online pharmacy PrEP or PEP service delivery via MYDAWA

Figure S2. Annual cost by categories (2023 USD)

Figure S3. Scenario analysis: estimated total and unit cost by client volume

Table S1. Cost categories, description, and data sources for online PrEP/PEP provision

| Cost Category | Type of Cost | Description | Data Source |
| --- | --- | --- | --- |
| Salaries and Benefits | Recurrent | Includes salaries and benefits (e.g., insurance) for personnel supporting online PrEP/PEP provision. | Project budgets and expense reports |
| Microplanning and Training | Start-up & Recurrent | 1) Planning activities for project implementation during the start-up period including stakeholder meetings and sensitization meetings. 2) Expenses for conducting training workshops during the start-up period and refresh trainings for project staff at MYDAWA. | Project budgets, expense reports, meeting agenda, and monitoring forms |
| Demand Generation and Marketing | Start-up & Recurrent | Includes the expenses for conducting online and in-person demand generation activities and the cost of printed posters and flyers. | Project budgets and expense reports |
| Clinical Consultation | Recurrent | Includes the value of time of study personnel to conduct clinical consultation during online PrEP/PEP visits. | Project budgets, expense reports, time-motion observations |
| Delivery of HIVST and Drugs | Recurrent | 1) Includes the value of time of warehouse staff and pharmtech riders to deliver HIVST and drugs. 2) Commodity cost of HIVST kit. | Project budgets, expense reports, time-motion observations |
| User Support | Recurrent | Includes the value of time of call center staff to provide user support to PrEP/PEP users. | Project budgets, expense reports, and phone call logs |
| Equipment, System Development, Utilities, and Fuel | Start-up & Recurrent | 1) Resources and inputs to design the Chabot solution, call center solution, telehealth clinical consultation solution, and AI system to read HIVST images. 2) Includes licensing and maintenance cost of the systems and utilities (internet, SMS, and electricity). 3) Investments that last longer than 1 year, including laptops, furniture, and the clinic space. | Project budgets, expense reports, and invoices. |
| Drug Costs (Economic Costs) | Recurrent | Includes the cost of PrEP drug per 30-day supply, the cost of PEP drug per month, and distribution and storage cost. | CHAI’s ARV Benchmark Price |
| Vehicles (Economic Costs) | Start-up | Motorbikes used by pharmtech staff to deliver HIVST and medications. | Staff interview |

Table S2. Input costs of key PrEP/PEP delivery components (2023 USD)

| Item | Cost (2023 USD) | Source |
| --- | --- | --- |
| PrEP drug per 30-day supply* | 4.28 | CHAI |
| PEP drug per month^†^ | 6.03 | CHAI |
| HIVST kit | 2 | MYDAWA budget |
| Subsidisation of SureCheck | 4.5 | MYDAWA budget |
| Subsidisation of Mylan | 3.5 | MYDAWA budget |
| Fuel allowance per year^‡^ | 7.45 | MYDAWA budget |
| Bike allowance per year^‡^ | 7.45 | MYDAWA budget |
| License and riding allowance per year^‡^ | 9.93 | MYDAWA budget |
| Boda (economic cost only) | 19 | MYDAWA budget |

^*^Acriptega includes 3 active ingredients: Dolutegravir (DTG), Lamivudine (3TC), Tenofovir (TDF) with the following strengths: Dolutegravir: 50mg; Lamivudine: 300mg; Tenofovir disoproxil: 300mg.

^†^Truvada contains 200 mg of emtricitabine (FTC) and 300 mg of tenofovir disoproxil fumarate (TDF).

^‡^Resources were allocated to online PrEP/PEP services based on the proportion of time dedicated to PrEP/PEP delivery.

Table S3. List of key model assumptions

1. Recurrent demand generation activities occur once every year to keep up with current trends.

2. All staff work 8 hours every day or 40 hours per week.

3. All deliveries are single delivery (not in batch) and one-step delivery (HIVST+drugs) for PrEP/PEP purchases in the base case.

4. Assume half of permanent staff have one child and one partner covered by health insurance.

5. Assume half of call center staff are permanent staff and the other half are contract staff.

6. Assume each client uploads 1 image for obtaining HIVST result.

7. Assume a new/unsupported HIVST kit that requires development of AI algorithms occurs once every ten years.

8. Assume all employees at MYDAWA are hired locally and no need for visa/international fees.

9. Assume 20% of the MYDAWA clinic's usage was allocated to the online PrEP/PEP provisions.

10. Assume the cost of medication purchase from the manufacturer as well as storage and transportation costs is charged at 8% of the cost of the product (Roberts, 2019).

11. Assume that all prescriptions or clinical consultations happen online.

12. 10% of PEP clients come in for their follow up visit. Around 80% will text the clinical officers to say that they took an HIV test and are negative and the remaining 10% are lost to follow up.

13. In the base case, assume no AI system is used for interpreting HIVST result. We have added 5 minutes to the observed CO time per client encounter with HIV test to account for time saved by the AI providing cleaned images (PEP initiation visit, PrEP initiation visit, PrEP continuation visit). We have decreased the % time for the Systems Administrator by 10% to account for less troubleshooting and ongoing IT support needed without AI. We have decreased the % time for the Technical Project Manager by 25% to account for less troubleshooting and ongoing IT support needed without AI

We have increased the cost of HIV test delivery by 7.5% to account for additional HIV tests that would need to be provided due to unreadable images in the absence of AI

Table S4. Scenario analysis of using $1 HIVST kit

| Cost Category | Total Cost: PrEP (n=229) | | Total Cost: PEP (n=1320) | | Unit Cost: PrEP (n=229) | | Unit Cost: PEP (n=1320) | |
| --- | --- | --- | --- | --- | --- | --- | --- | --- |
|  | Economic | Financial | Economic | Financial | Economic | Financial | Economic | Financial |
| Salaries and Benefits | $1,138 | $1,138 | $6,559 | $6,559 | $5.0 | $5.0 | $5.0 | $5.0 |
| Microplanning and Training | $277 | $277 | $1,596 | $1,596 | $1.2 | $1.2 | $1.2 | $1.2 |
| Demand Generation and Marketing | $5,034 | $5,034 | $29,017 | $29,017 | $22.0 | $22.0 | $22.0 | $22.0 |
| Clinical Consultation | $1,800 | $1,800 | $8,832 | $8,832 | $7.9 | $7.9 | $6.7 | $6.7 |
| Delivery of HIVST and Drugs | $9,454 | $9,454 | $35,176 | $35,176 | $41.3 | $41.3 | $26.6 | $26.6 |
| User Support | $104 | $104 | $599 | $599 | $0.5 | $0.5 | $0.5 | $0.5 |
| Equipment, System Development, Utilities, and Fuel | $1,259 | $1,259 | $7,258 | $7,258 | $5.5 | $5.5 | $5.5 | $5.5 |
| Drug Costs (Economic Costs) | $11,753 | $0 | $7,962 | $0 | $51.3 | $0.0 | $6.0 | $0.0 |
| Vehicles (Economic Costs) | $3 | $0 | $16 | $0 | $0.01 | $0.0 | $0.01 | $0.0 |
| Total Cost | $30,821 | $19,066 | $97,015 | $89,037 |  |  |  |  |
| Cost Per Client | $134.6 | $83.3 | $73.5 | $67.5 |  |  |  |  |
| Cost per PrEP Client Month | $60.8 | $37.6 |  |  |  |  |  |  |

Table S5. Scenario analysis of including AI HIV testing system

| Cost Category | Total Cost: PrEP (n=229) | | Total Cost: PEP (n=1320) | | Unit Cost: PrEP (n=229) | | Unit Cost: PEP (n=1320) | |
| --- | --- | --- | --- | --- | --- | --- | --- | --- |
|  | Economic | Financial | Economic | Financial | Economic | Financial | Economic | Financial |
| Salaries and Benefits | $1,226 | $1,226 | $7,065 | $7,065 | $5.4 | $5.4 | $5.4 | $5.4 |
| Microplanning and Training | $277 | $277 | $1,596 | $1,596 | $1.2 | $1.2 | $1.2 | $1.2 |
| Demand Generation and Marketing | $5,034 | $5,034 | $29,017 | $29,017 | $22.0 | $22.0 | $22.0 | $22.0 |
| Clinical Consultation | $1,299 | $1,299 | $7,007 | $7,007 | $5.7 | $5.7 | $5.3 | $5.3 |
| Delivery of HIVST and Drugs | $9,157 | $9,157 | $36,628 | $36,628 | $40.0 | $40.0 | $27.7 | $27.7 |
| User Support | $104 | $104 | $599 | $599 | $0.5 | $0.5 | $0.5 | $0.5 |
| Equipment, System Development, Utilities, and Fuel | $3,893 | $3,893 | $22,437 | $22,437 | $17.0 | $17.0 | $17.0 | $17.0 |
| Drug Costs (Economic Costs) | $11,753 | $0 | $7,962 | $0 | $51.3 | $0.0 | $6.0 | $0.0 |
| Vehicles (Economic Costs) | $3 | $0 | $16 | $0 | $0.01 | $0.0 | $0.01 | $0.0 |
| Total Cost | $32,744 | $20,989 | $112,328 | $104,350 |  |  |  |  |
| Cost Per Client | $143.0 | $91.7 | $85.1 | $79.1 |  |  |  |  |
| Cost per PrEP Client Month | $64.6 | $41.4 |  |  |  |  |  |  |

Assumptions:

1. We have included the cost of AI development
2. We have included the cost of AI system integration and set up
3. We have included the per-image cost of AI interpretation/image cleaning
4. We have decreased 5 minutes to CO time per client encounter with HIV test to account for time saved by the AI providing cleaned images (PEP initiation visit, PrEP initiation visit, PrEP continuation visit)
5. We have increased the % time for the Systems Administrator by 10% to account for troubleshooting and ongoing IT support needed with AI
6. We have increased the % time for the Technical Project Manager by 25% to account for troubleshooting and ongoing IT support needed with AI
7. We have decreased the cost of HIV test delivery by 7.5% to account for HIV tests that would need to be provided due to unreadable images in the absence of AI

Table S6. Scenario analysis of varying duration of PrEP initiation and continuation visits

|  | Duration + 30% | | Duration - 30% | |
| --- | --- | --- | --- | --- |
|  | **Economic** | **Financial** | **Economic** | **Financial** |
| Total Cost | $31,752 | $19,996 | $30,671 | $18,916 |
| Cost Per Client | $138.7 | $87.3 | $133.9 | $82.6 |
| Cost per PrEP Client Month | $62.6 | $39 | $60.5 | $37 |
| Cost per PrEP initiation | $76.6 | $44 | $73.9 | $42 |
| Cost per PrEP continuation | $59.6 | $27.2 | $58.3 | $25.9 |

Table S7. Scenario analysis of using lower-cadre pharmtech workers

| Cost Category | Total Cost: PrEP (n=229) | | Total Cost: PEP (n=1320) | | | Unit Cost: PrEP (n=229) | | | Unit Cost: PEP (n=1320) | | |
| --- | --- | --- | --- | --- | --- | --- | --- | --- | --- | --- | --- |
|  | Economic | Financial | | Economic | Financial | | Economic | Financial | | Economic | Financial |
| Salaries and Benefits | $1,138 | $1,138 | | $6,559 | $6,559 | | $5.0 | $5.0 | | $5.0 | $5.0 |
| Microplanning and Training | $277 | $277 | | $1,596 | $1,596 | | $1.2 | $1.2 | | $1.2 | $1.2 |
| Demand Generation and Marketing | $5,034 | $5,034 | | $29,017 | $29,017 | | $22.0 | $22.0 | | $22.0 | $22.0 |
| Clinical Consultation | $1,800 | $1,800 | | $8,832 | $8,832 | | $7.9 | $7.9 | | $6.7 | $6.7 |
| Delivery of HIVST and Drugs | $7,484 | $7,484 | | $27,846 | $27,846 | | $32.7 | $32.7 | | $21.1 | $21.1 |
| User Support | $104 | $104 | | $599 | $599 | | $0.5 | $0.5 | | $0.5 | $0.5 |
| Equipment, System Development, Utilities, and Fuel | $1,259 | $1,259 | | $7,258 | $7,258 | | $5.5 | $5.5 | | $5.5 | $5.5 |
| Drug Costs (Economic Costs) | $11,753 | $0 | | $7,962 | $0 | | $51.3 | $0.0 | | $6.0 | $0.0 |
| Vehicles (Economic Costs) | $3 | $0 | | $16 | $0 | | $0.01 | $0.0 | | $0.01 | $0.0 |
| Total Cost | $28,851 | $17,096 | | $89,684 | $81,706 | |  |  | |  |  |
| Cost Per Client | $126.0 | $74.7 | | $67.9 | $61.9 | |  |  | |  |  |
| Cost per PrEP Client Month | $56.9 | $33.7 | |  |  | |  |  | |  |  |

Figure S1. The proposed care pathway for online pharmacy PrEP or PEP service delivery via MYDAWA.

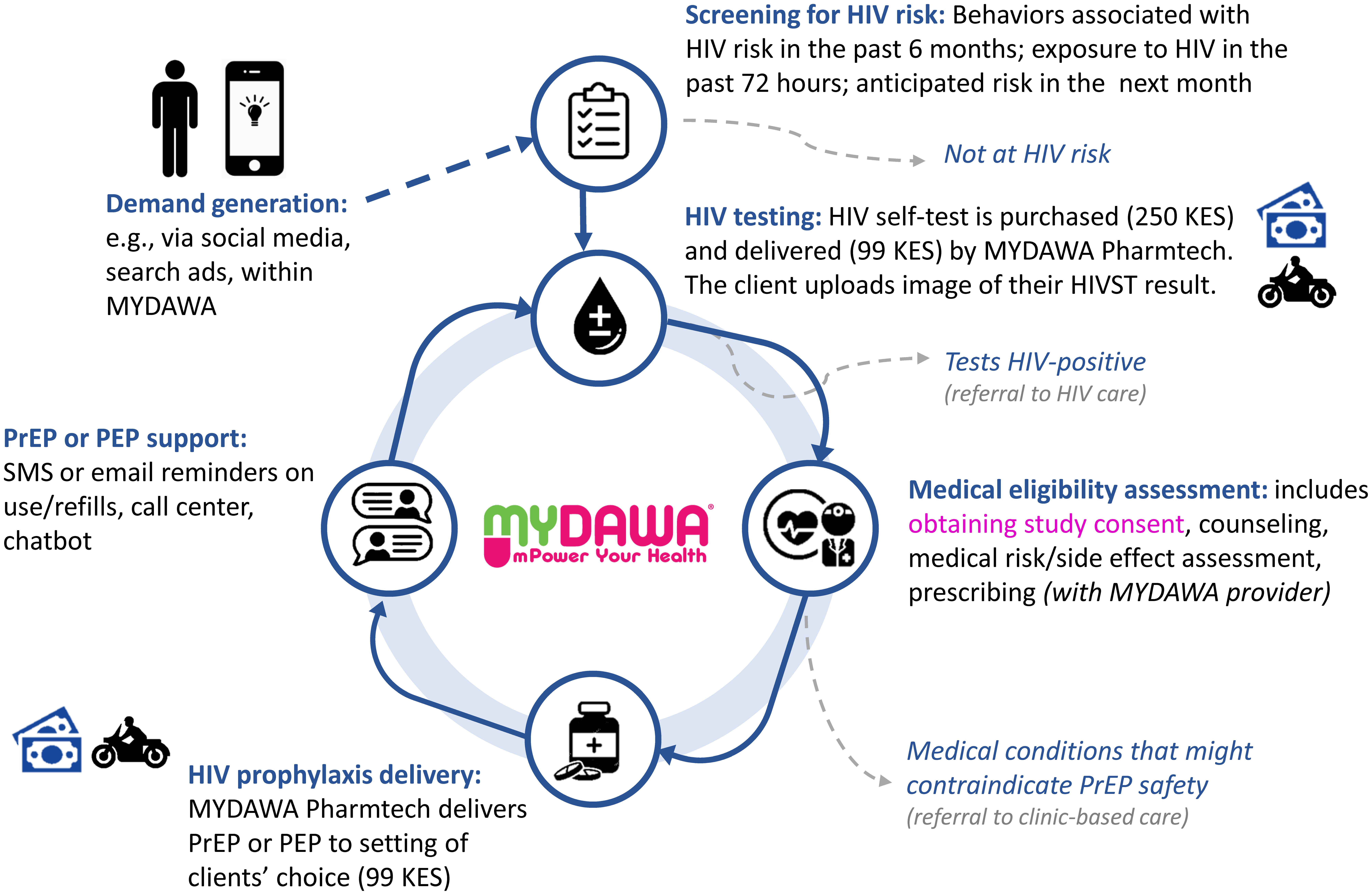

Figure S2. Annual cost by categories (2023 USD)

(A) PrEP provision

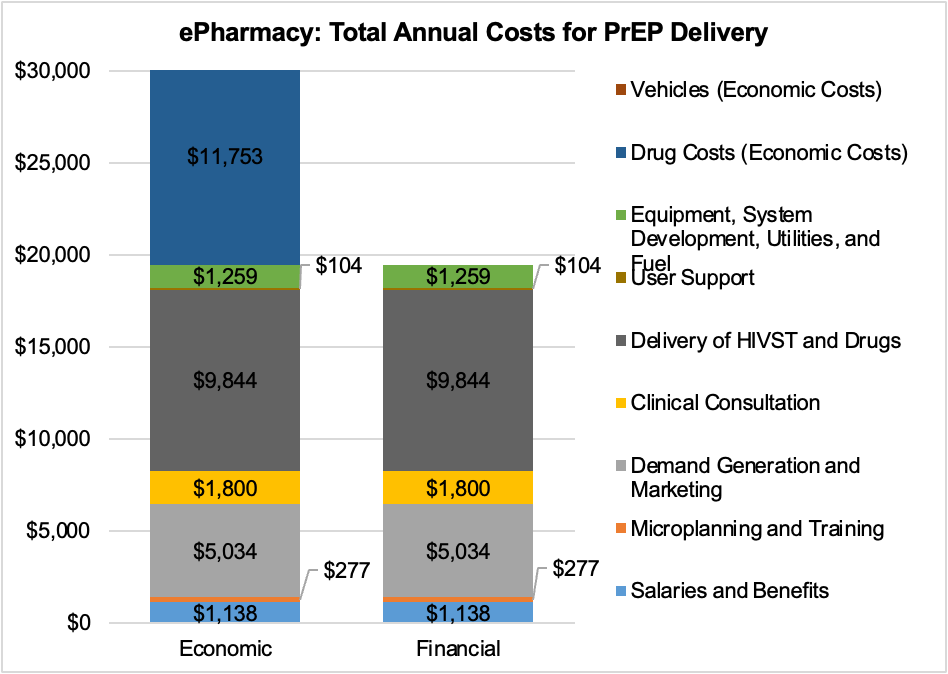

(B) PEP provision

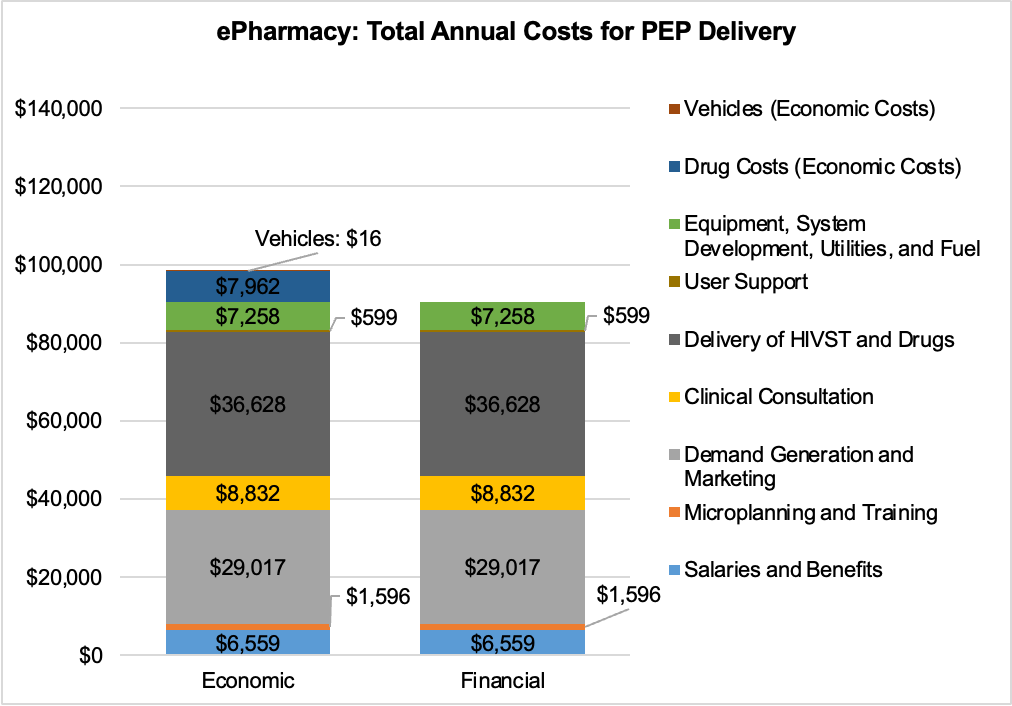

Figure S3. Scenario analysis: estimated total and unit cost by client volume

(A)

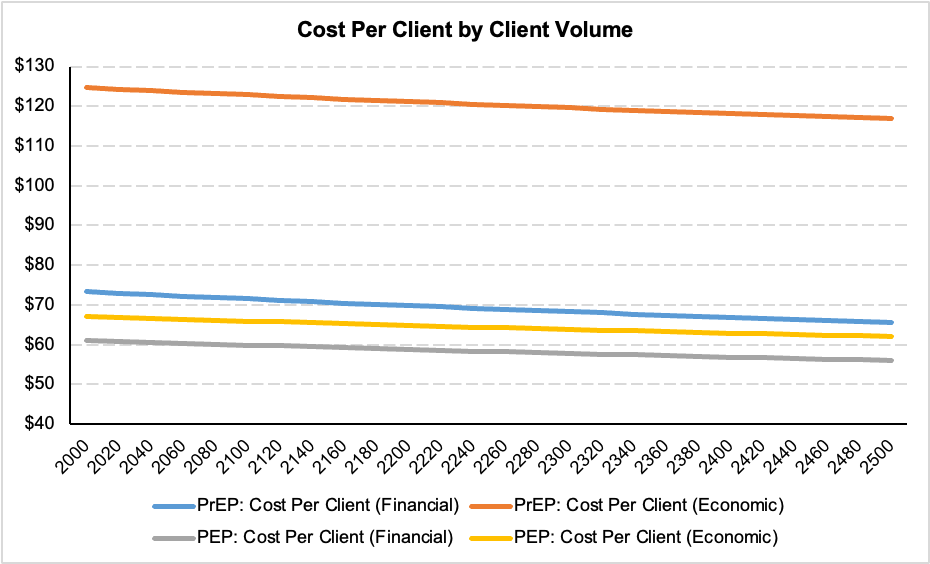

(B)

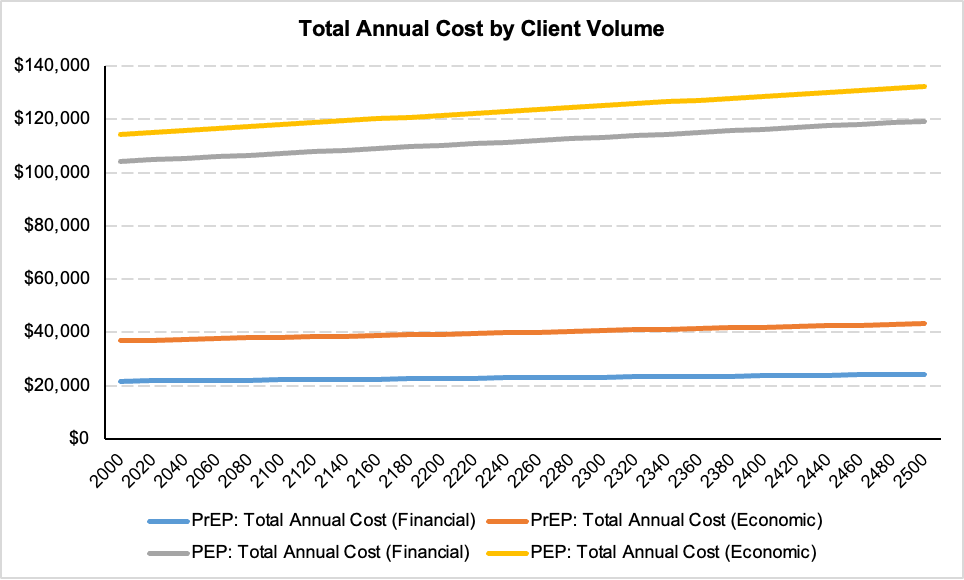
